# Living with Children and Adults’ Risk of COVID-19: Observational Study

**DOI:** 10.1101/2020.09.21.20196428

**Authors:** Rachael Wood, Emma C Thomson, Robert Galbraith, Ciara Gribben, David Caldwell, Jennifer Bishop, Martin Reid, Anoop S V Shah, Kate Templeton, David Goldberg, Chris Robertson, Sharon Hutchinson, Helen Colhoun, Paul McKeigue, David A McAllister

**Affiliations:** Usher Institute, University of Edinburgh, Edinburgh, UK; Public Health Scotland, Edinburgh, UK; MRC Centre for Virus Research, University of Glasgow, Glasgow, UK; Non-communicable Disease Epidemiology, London School of Hygiene and Tropical Medicine, London, UK; Department of cardiology, Imperial College NHS Trust, London, UK; School of Health and Life Sciences, Glasgow Caledonian University, Glasgow, UK; MRC Institute of Genetics & Molecular Medicine, University of Edinburgh, Edinburgh, UK; Institute of Health and Wellbeing, University of Glasgow, Glasgow, UK

## Abstract

**Objective:** Children are relatively protected from COVID-19, possibly due to cross-protective immunity. We investigated if contact with children also affords adults a degree of protection from COVID-19.

**Design:** Cohort study based on linked administrative data.

**Setting:** Scotland

**Study population:** All NHS Scotland healthcare workers and their household contacts as of March 2020.

**Main exposure:** Number of young children (0-11 years) living in the participant’s household.

**Main outcomes:** COVID-19 requiring hospitalisation, and any COVID-19 (any positive test for SARS-CoV-2) in adults aged ≥18 years between 1 March and 12 October 2020.

**Results:** 241,266, 41,198, 23,783 and 3,850 adults shared a household with 0, 1, 2, and 3 or more young children respectively. Over the study period, the risk of COVID-19 requiring hospitalisation was reduced progressively with increasing numbers of household children - fully adjusted hazard ratio (aHR) 0.93 per child (95% CI 0.79-1.10). The risk of any COVID-19 was similarly reduced, with the association being statistically significant (aHR per child 0.93; 95% CI 0.88-0.98). After schools reopened to all children in August 2020, no association was seen between exposure to young children and risk of any COVID-19 (aHR per child 1.03; 95% CI 0.92-1.14).

**Conclusion:** Between March and October 2020, living with young children was associated with an attenuated risk of any COVID-19 and COVID-19 requiring hospitalisation among adults living in healthcare worker households. There was no evidence that living with young children increased adults’ risk of COVID-19, including during the period after schools re-opened.

## Introduction

To date, children seem to be relatively protected from Severe Acute Respiratory Syndrome coronavirus 2 (SARS-CoV-2), being substantially less likely to develop symptomatic infection (COVID-19) or serious illness.[1,2] The underlying reasons are poorly understood, but differences in innate and/or acquired immune mechanisms may play a part.[3-11] Children may have increased innate immune responsiveness following vaccinations and high exposure to respiratory viruses.[12] Alternatively, pre-exposure to antigenically-similar infectious agents (providing subsequent specific cross-protection against SARS-CoV-2) may also be relevant. Children have higher levels of exposure to endemic coronaviruses than adults.[13,14] Evidence exists for B and T-cell cross-reactivity between SARS-CoV-2 and endemic coronaviruses [4–10]; and SARS-CoV-2 responsive T cells have been shown to provide protection against COVID-19[11].

Given this, we reasoned that adults who are close contacts of children might also be relatively protected from SARS-CoV-2 due to a degree of immune cross-protection. This could be relevant to decisions to close nurseries and schools in response to SARS-CoV-2[15] and teachers’ perceptions of workplace safety.[16] Since few relevant studies exist,[17] we used a recently-reported cohort of around 160,000 healthcare workers and 250,000 household members in Scotland,[18] to test the hypothesis that risk of COVID-19 in adults is attenuated among those living with young children.

## Methods

### Population, data sources, and record linkage

Full details of the population studied are reported elsewhere.[18] In brief, we identified healthcare workers in Scotland (aged 18 to 65) using databases including all individuals directly employed by the NHS, and all General Practitioners providing services to the NHS, as of March 2020. We identified other members of the healthcare workers’ households (all ages) using the NHS Scotland master patient index and exact address matching. We linked these data to multiple Scotland-wide databases indicating virology testing for SARS-CoV-2, hospitalisation, critical care admission, and death. Exposure, covariate, and outcome data were examined for all adults aged ≥18 living in a healthcare worker household. All data from 1 March through 12 October 2020 were included.

### Outcome

The primary outcome was COVID-19 requiring hospitalisation, defined as a first positive PCR test[19] for SARS-CoV-2 up to 28 days prior to, or during, a hospital admission. Secondary outcomes were any COVID-19 (defined as any positive test for SARS-CoV-2), and severe COVID-19 (defined as a positive test for SARS-CoV-2 up to 28 days prior to admission for critical care or death).

### Exposure

The primary exposure was the number of young children (aged 0 to 11) in each household. Additional analyses examined risk by the number of pre-school children (aged 0-4), primary school children (aged 5-11), older children (aged 12-17), and other adults (aged ≥18).

### Covariates

Data on age, sex, and deprivation (Scottish Index of Multiple Deprivation (SIMD) quintile) were obtained from the linked databases. Pre-specified comorbidities (see Table S1) were defined using previous hospitalisation and prescribing data. Ethnicity was imputed from forename and surname using the ONOMAP algorithm[20].

Occupational covariates were defined at the household level based on the characteristics of the member who was a healthcare worker. These included the healthcare worker’s occupation (e.g. medical, nursing), potential exposure to SARS-CoV-2 (e.g. patient-facing role or not), seniority, length of service, immigration status, and full/part-time working status. In households with more than one healthcare worker, the highest risk category was applied. A causal diagram showing the assumed relation between these covariates is provided in Figure S1.

### Statistical analysis

We plotted the cumulative incidence of hospitalisation for COVID-19 among adults according to the number of young children in each household. We modelled COVID-19 requiring hospitalisation, any COVID-19, and severe COVID-19 using Cox regression, calculating robust standard errors to allow for clustering due to shared household membership and stratifying on groups of health board areas to allow for differences in baseline hazard. We present effect estimates for minimal models adjusting for age, full models including all covariates, and intermediate models. We conducted a range of sensitivity analyses incorporating additional covariates, and/or restricting the population examined.

### Ethics

This project was approved by the Public Benefit and Privacy Panel (2021-0013).

## Results

Of the 310,097 adults living in a healthcare worker household, 241,266 (78%), 41,198 (13%), 23,783 (7.8%) and 3,850 (1.2%) shared their household with 0, 1, 2 and 3 or more young children respectively. Compared to adults living with no young children, those living with children were on average 5 years younger; were less likely to live in the most deprived areas; were more likely to be of non-white ethnicity, and were less likely to have comorbidities (Table 1 and Supplementary Table S1). Adults living with young children were slightly more likely to be tested for SARS-CoV-2 (Table 1).

**Table 1.** Characteristics of adult household contacts by number of young children.

|  | 0 children<br>aged 0-11 | 1 child<br>aged 0-11 | 2 children<br>aged 0-11 | 3+ children aged<br>0-11 |
| --- | --- | --- | --- | --- |
| **Number of adults** | 241266 | 41198 | 23783 | 3850 |
| **Adults who are healthcare workers** | 121004 (50.15) | 22025 (53.46) | 13179 (55.41) | 2237 (58.10) |
| **Age, mean (standard deviation)** | 44.53 (15.04) | 39.82 (10.58) | 39.01 (8.08) | 38.47 (6.62) |
| **Male** | 105116 (43.57) | 17639 (42.82) | 10803 (45.42) | 1783 (46.31) |
| **Scottish index of multiple deprivation** |  |  |  |  |
| *1 - most deprived* | 37242 (15.44) | 5655 (13.73) | 2447 (10.29) | 373 (9.69) |
| *2* | 46147 (19.13) | 7700 (18.69) | 3599 (15.13) | 582 (15.12) |
| *3* | 48659 (20.17) | 7491 (18.18) | 4280 (18.00) | 706 (18.34) |
| *4* | 52847 (21.90) | 9701 (23.55) | 6102 (25.66) | 999 (25.95) |
| *5 - least deprived* | 56371 (23.36) | 10651 (25.85) | 7355 (30.93) | 1190 (30.91) |
| **Race/ethnicity - Non-white** | 7768 (3.22) | 2088 (5.07) | 1162 (4.89) | 259 (6.73) |
| **Comorbidity count** |  |  |  |  |
| *None* | 207796 (86.13) | 37315 (90.57) | 21924 (92.18) | 3564 (92.57) |
| *One* | 24897 (10.32) | 3231 (7.84) | 1579 (6.64) | 252 (6.55) |
| *Two or more* | 8573 (3.55) | 652 (1.58) | 280 (1.18) | 34 (0.88) |
| **Occupation of healthcare worker in household** |  |  |  |  |
| *Nursing and midwifery* | 102514 (42.49) | 18688 (45.36) | 10085 (42.40) | 1530 (39.74) |
| *Administrative services* | 44929 (18.62) | 6710 (16.29) | 3236 (13.61) | 404 (10.49) |
| *Support services* | 27294 (11.31) | 3232 (7.85) | 1386 (5.83) | 236 (6.13) |
| *Medical and dental* | 20836 (8.64) | 4326 (10.50) | 3586 (15.08) | 849 (22.05) |
| *Allied health profession* | 20007 (8.29) | 3798 (9.22) | 2974 (12.50) | 442 (11.48) |
| *Other* | 25686 (10.65) | 4444 (10.79) | 2516 (10.58) | 389 (10.10) |
| **Occupational role of healthcare worker in household** |  |  |  |  |
| *Non-patient facing* | 50441 (20.91) | 7453 (18.09) | 3667 (15.42) | 471 (12.23) |
| *Patient facing* | 137697 (57.07) | 25461 (61.80) | 15485 (65.11) | 2627 (68.23) |
| *Undetermined* | 53128 (22.02) | 8284 (20.11) | 4631 (19.47) | 752 (19.53) |
| **Part time working in healthcare worker in household** |  |  |  |  |
| *Whole time* | 147608 (61.18) | 19482 (47.29) | 8296 (34.88) | 1206 (31.32) |
| *Part time* | 88351 (36.62) | 20329 (49.34) | 14183 (59.64) | 2281 (59.25) |
| *Not recorded* | 5307 (2.20) | 1387 (3.37) | 1304 (5.48) | 363 (9.43) |
| **Tested for SARS-CoV-2** | 14736 (6.11) | 2835 (6.88) | 1823 (7.67) | 354 (9.19) |

Statistics are the number (percentage) of adults with each characteristic except for age, which is given as the mean and standard deviation.

Household composition differed according to the number of young children. Households with more children were more likely to include 2 or more adults. More than a quarter of adults who shared a household with a single child under 11 also shared a household with a child aged 12-17 (Supplementary Table S2).

### COVID-19 requiring hospitalisation

Compared to adults living with no young children, the risk of COVID-19 requiring hospitalisation was reduced in those living with children (Figure 1). The unadjusted hazard ratio (HR) for COVID-19 requiring hospitalisation was 0.77 per each additional young child in the household (95% CI 0.65-0.90, p 0.001, Table 2). On adjusting for adults’ age, this association was attenuated (HR per child 0.88; 95% CI 0.75-1.04). Further smaller changes were seen after adjusting for other potential confounders (sex, deprivation, occupation, professional role, staff/non-staff status, length of service, number of adolescents and adults in household, comorbidity count plus selected comorbidities - see Table S1 - and full/part time working status), with the fully adjusted HR (aHR) being 0.93 per child (95% CI 0.79-1.10).

**Table 2.** Risk and hazard ratios for COVID-19 requiring hospitalisation for adults living in healthcare worker households by number of young children.

|  | No children aged 0-11 | 1 child aged 0-11 | 2 children aged 0-11 | 3+ children aged 0-11 | Per child |
| --- | --- | --- | --- | --- | --- |
| N adults with COVID-19 requiring hospitalisation | 390 | 59 | 20 | 2 | - |
| Total N adults | 241266 | 41198 | 23783 | 3850 | - |
| Risk per 10,000 | 16.2 | 14.3 | 8.4 | 5.2 | - |
| Unadjusted | 1 | 0.89 (0.67-1.17) | 0.53 (0.34-0.83) | 0.33 (0.08-1.32) | 0.77 (0.65-0.90) |
| Model 1 | 1 | 1.09 (0.82-1.46) | 0.69 (0.44-1.10) | 0.44 (0.11-1.79) | 0.88 (0.75-1.04) |
| Model 2 | 1 | 1.11 (0.83-1.49) | 0.72 (0.45-1.15) | 0.44 (0.11-1.78) | 0.90 (0.76-1.05) |
| Model 3 | 1 | 1.13 (0.84-1.52) | 0.73 (0.46-1.16) | 0.45 (0.11-1.81) | 0.90 (0.77-1.06) |
| Model 4 | 1 | 1.17 (0.87-1.57) | 0.78 (0.49-1.26) | 0.49 (0.12-1.97) | 0.93 (0.79-1.10) |

Hazard ratios obtained from Cox proportional hazard models. Model 1 adjusts for adults' age using a penalised spline function. Model 2 additionally adjusts for sex, Scottish Index of Multiple Deprivation quintile, occupation (eg nursing, medical), occupational role (patient facing, non-patient facing, undetermined), healthcare worker (yes/no), length of service, number of children aged 12 to 17 in household, number of adults in household. Model 3 additionally adjusts for the comorbidity count and specific conditions (ischaemic heart disease, other heart disease, other circulatory system diseases, advanced chronic kidney disease, asthma and chronic lower respiratory disease, neurological disorders, decompensated liver disease, any immunological condition, malignant neoplasms, disorders of oesophagus, stomach and duodenum, type 1 diabetes and type 2 diabetes). Model 4 additionally adjusts for part-time status (additional model not pre-specified).

**Figure 1.**
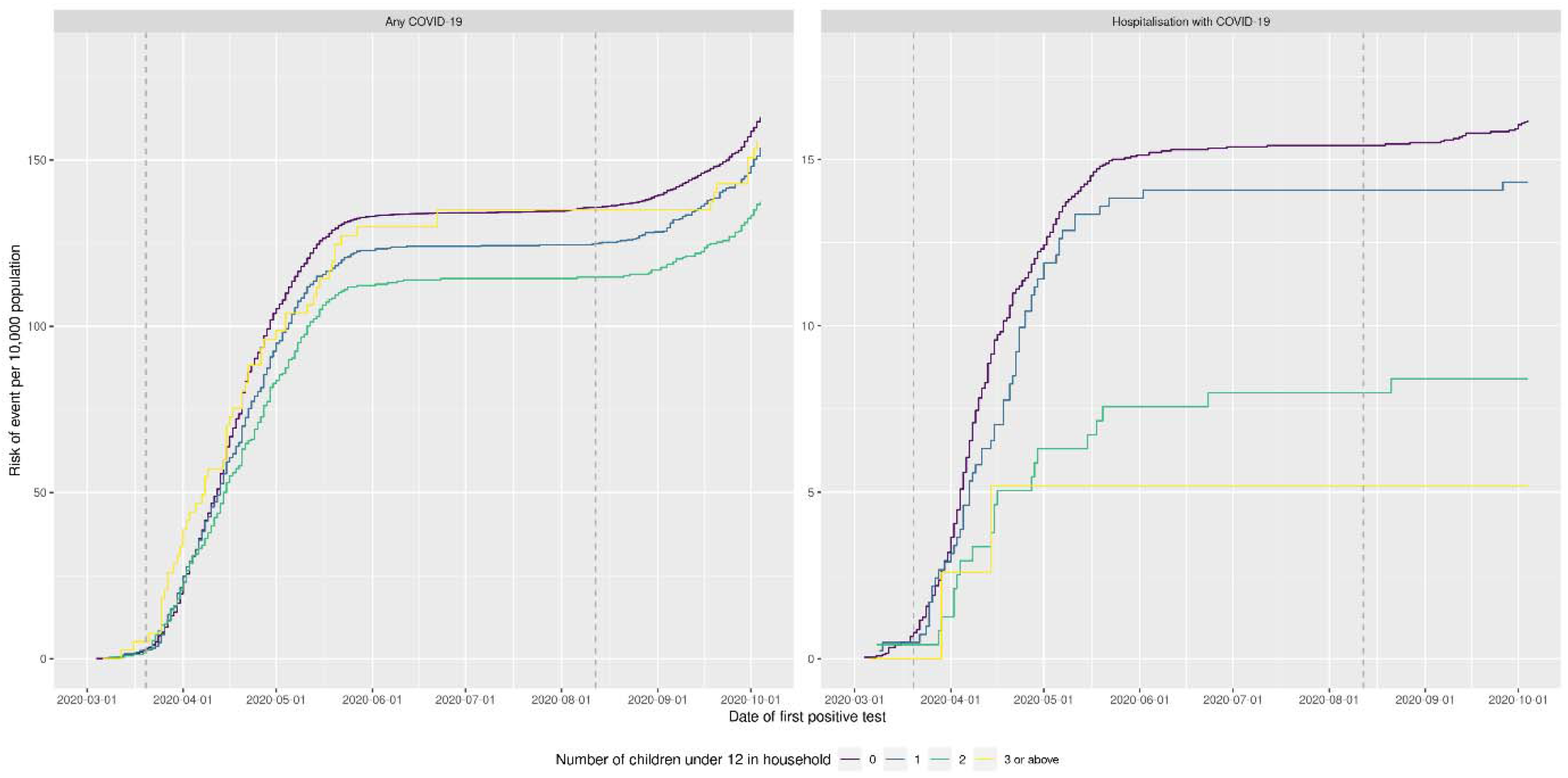
Risk of any COVID-19 and of COVID-19 requiring hospitalisation in adults living in healthcare worker households by number of young children (aged 0 to 11) Cumulative incidence (risk) plots of any COVID-19 and of COVID-19 requiring hospitalisation by number of young children (aged 0 to 11) in household. Vertical dotted lines indicate dates when schools closed and re-opened.

Similar associations were found when analysis was restricted to households where at least one adult was a patient-facing healthcare worker (aHR per child 0.89; 95% CI 0.74-1.08, Supplementary Table S3). In a stratified analysis, the aHR per child was 1.11 (95% CI 0.92-1.35) for adults living in households containing a full-time healthcare worker and 0.65 (95% CI 0.46-0.91) for those in households containing a part time healthcare worker (p-interaction 0.002). A further stratified analysis found similar results for adults with (aHR 0.91; 95% CI 0.75-1.10) and without (aHR 0.90; 95% CI 0.65-1.25) any comorbidities.

### Any COVID-19

Compared to adults living with no young children, the risk of any COVID-19 was reduced in those living with children (Figure 1). In the full study population, the fully aHR for any COVID-19 was similar to that seen for COVID-19 requiring hospitalisation, but with narrower confidence intervals (aHR per child 0.93 95% CI 0.88-0.98, Table 3). The inverse association between number of young children in the household and risk of any COVID-19 was similar in adults with (aHR per child 0.84; 95% CI 0.72-0.99) and without (aHR 0.93; 95% CI 0.88-0.97) comorbidities, and in adults living in households containing a full (aHR 0.95; 95% CI 0.88-1.02) and part time (aHR 0.88; 95% CI 0.81-0.94) healthcare worker (p-interaction 0.44).

**Table 3.**
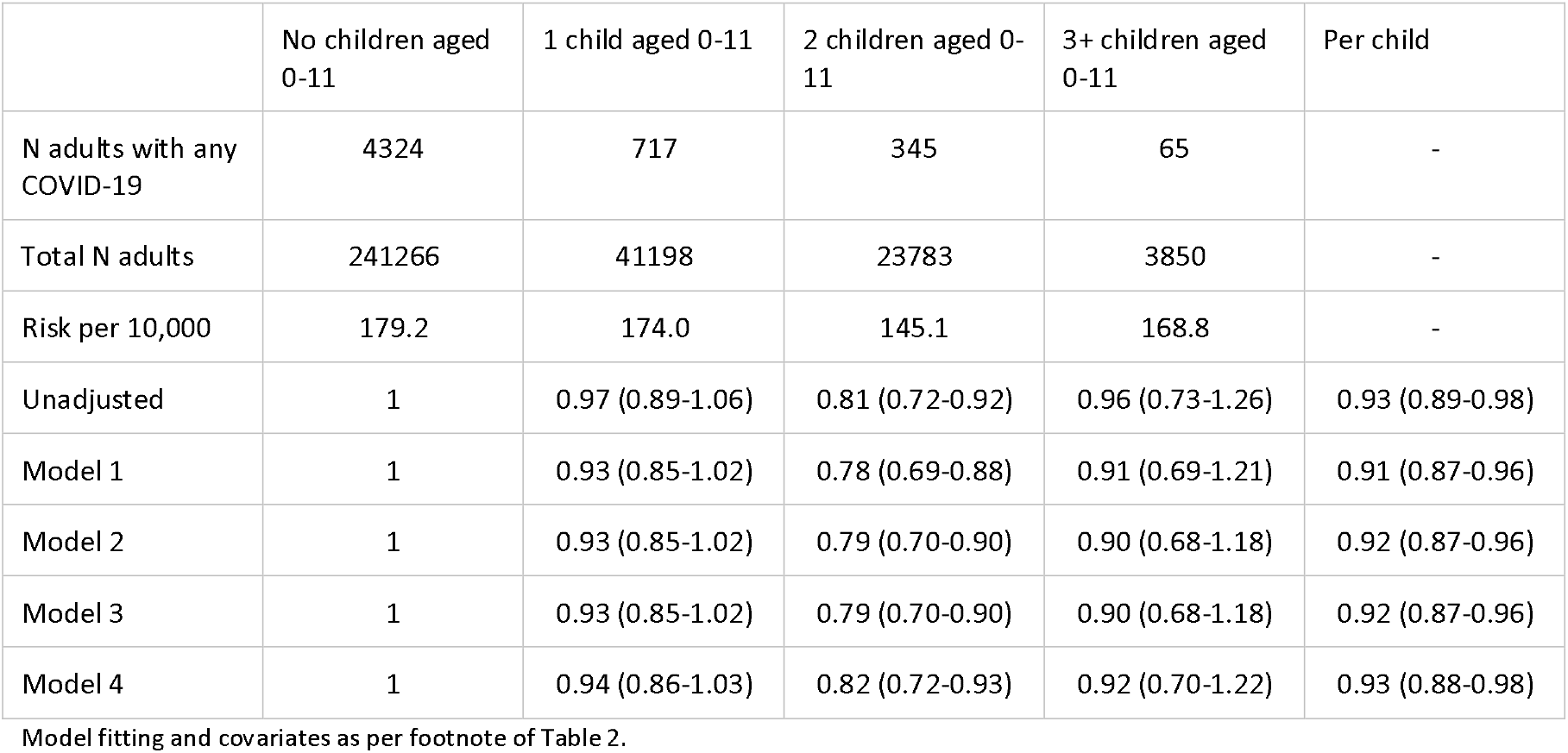
Risk and hazard ratios for any COVID-19 for adults living in healthcare worker households by number of young children.

Further analysis of the risk of any COVID-19 revealed stronger inverse associations for adults living with pre-school children, compared to those living with primary school children, adolescents, or other adults (Table 4). Similar differences between the age-groups, but with wider 95% confidence intervals reflecting the smaller numbers of events, were also found for the primary outcome of COVID-19 requiring hospitalisation (Supplementary Table S4).

**Table 4.** Hazard ratios for any COVID-19 for adults living in healthcare worker households by number of persons of different ages.

|  | Per child<br>aged 0 to 4 | Per child<br>aged 5 to 11 | Per child<br>aged 12 to 17 | Per adult<br>aged 18 or above |
| --- | --- | --- | --- | --- |
| Unadjusted | 0.88 (0.81-0.96) | 1.07 (1.01-1.13) | 0.87 (0.84-0.89) | 0.97 (0.91-1.02) |
| Model 1 | 0.87 (0.80-0.95) | 1.03 (0.97-1.09) | 0.88 (0.85-0.90) | 0.94 (0.88-0.99) |
| Model 2 | 0.86 (0.78-0.94) | 1.05 (0.99-1.12) | 1.04 (1.02-1.07) | 0.95 (0.89-1.01) |
| Model 3 | 0.86 (0.78-0.94) | 1.05 (0.99-1.12) | 1.04 (1.02-1.07) | 0.95 (0.89-1.01) |
| Model 4 | 0.87 (0.80-0.95) | 1.06 (1.00-1.12) | 1.04 (1.01-1.07) | 0.96 (0.90-1.02) |

### School re-opening

Nurseries and schools in Scotland were closed from 23 March 2020 until the end of the school year in late June 2020. They reopened to all children on 12 August 2020 and remained open until the Christmas holiday. Some childcare or school provision was available for children of key workers, including healthcare workers, between March and August. Between 12 August and 12 October 2020 (the end date of our study period), there were an additional 1,337 cases of any COVID and 20 cases of COVID-19 requiring hospitalisation among the adults in our study population (Figure 1) and the aHR for any COVID-19 was 1.03 (95% CI 0.92-1.14) per each additional young child in the household.

### Additional analyses

Results for the much less common outcome, severe COVID-19, are shown in supplementary Tables S5 and S6. Restricting the analysis to individuals identified via the ONOMAP algorithm as white did not modify the associations for any of the outcomes. The full set of regression coefficients and standard errors for all fitted models are provided at https://github.com/ChronicDiseaseEpi/hcw.

## Discussion

We examined the hypothesis that the risk of COVID-19 in adults is attenuated for those with high exposure to young children (0-11 years) due to presumed cross-protective immunity. Among a cohort of over 300,000 adults living in a household with a healthcare worker in Scotland, the risk of testing positive for SARS-CoV-2 over the period March to October 2020 was slightly lower for individuals living with young children, and this reduction persisted after adjusting for potential confounders. The risk of COVID-19 requiring hospitalisation (primary outcome specified) was also lower for those living with young children, although this finding did not reach statistical significance.

Very few studies have directly examined whether contact with children affords adults protection from SARS-CoV-2. Prior to our recent preprint,[21] only one study touching on this question was identified.[17] In this German study, 1,186 of 4,010 patients who had recovered from COVID-19 responded to a survey. The proportion of individuals reporting regular contact with children aged <11 years was lower than expected based on general population rates. More recently a preprint has been published by the OpenSAFELY group.[22] In this general population sample of 12 million people in England, between February and July 2020, adults sharing a household with children aged 0-11 years were not at increased risk of any COVID-19 or COVID-19 requiring hospitalisation or critical care admission, and were at a lower risk of death from COVID-19. These studies are congruent with our findings and together suggest that, to date, exposure to young children is not associated with increased risk of COVID-19 in adults, but with a small reduction in risk.

The risk to adults of COVID-19 will presumably reflect a dynamic balance between the risk that children may directly transmit SARS-CoV-2 to their adult contacts, and the possibility that they may enhance cross-protective immunity through prior transmission of other respiratory viruses. In this context, our current study provides reassurance that adults sharing a household with young children remained at no increased risk of COVID-19 during August to October 2020 when schools were reopened and community transmission of SARS-CoV-2 was occurring.

This study has some limitations. The observed inverse association between living with young children and adults’ risk of COVID-19 was not strong, and could be a chance finding. Our primary outcome, COVID-19 requiring hospitalisation, was uncommon; hence HR confidence intervals were wide. Although we suspected that statistical power would be limited, we pre-specified this as the primary outcome as we were concerned that high rates of (non-SARS-CoV-2) acute respiratory infection in households with small children might have led to higher levels of testing for SARS-CoV-2 and hence biased ascertainment of any COVID-19. The level of testing was indeed higher among those adults who shared a household with young children. However, point estimates for COVID-19 requiring hospitalisation and any COVID-19 were similar, and for the latter they were statistically significant.

Another possibility is that, despite extensive adjustment for potential confounders, the observed inverse association may be the result of residual confounding. On stratified analyses, the inverse association was evident for adults living in part time healthcare worker households, but less obvious for those living in full time healthcare worker households. Since part-time workers with increasing numbers of children likely work fewer hours (and therefore have lower occupational exposure to SARS-CoV-2), and since we lacked accurate data on hours worked during the pandemic, we cannot exclude the possibility of unmeasured confounding as a cause of the observed associations.

It has been suggested that the inverse association between living with young children and risk of COVID-19 may result from adults living with children spending less time outside the home in settings in which SARS-CoV-2 may be transmitted. While plausible, we are unable to find any empirical evidence in support of this view. For much of the pandemic across much of Scotland many social venues have been closed. In addition, evidence from the 2018 Scottish Household Survey (Supplementary Tables S7-10) suggests that adults who live with young children are as likely to visit restaurants and gyms as other adults of the same age, and indeed more likely to visit places of worship and cinemas.

If a protective effect of children on COVID-19 rate and severity in their adult contacts is confirmed, this could involve cross-reactive immunity to endemic coronavirus infections acquired outside the home, e.g. at nursery or school. Firstly, evidence of antigenic similarity between N proteins of SARS-CoV-2 and those of endemic beta coronaviruses (strains Cov-OC43 and Cov-NL63) has now been shown in studies of cell-mediated immunity. There is also evidence of cross-reactivity in antibody-mediated immunity, although it is uncertain how well this protects against COVID-19.[4–10] Secondly, respiratory samples obtained from children during investigation for respiratory tract infections show high levels of seasonal, endemic coronaviruses,[13,14] Thirdly, as well as having higher rates of exposure to such viruses, children may transmit seasonal coronaviruses to their household contacts. Younger adults (aged 15-44) that include those most likely to live with young children, have higher levels of antibodies to N proteins of CoV-OC43 than do older adults,[23] although whether this reflects exposure at home via contact with children, or elsewhere, is unknown. It would be important to compare the prevalence of antibodies to SARS-CoV-2, and to seasonal coronaviruses, in those with and without substantive exposure to children of different age groups.

Notwithstanding possible mechanisms, our findings provide sufficient evidence of a potentially interesting protective effect against COVID-19 for adults living with young children to warrant further study in other populations (e.g. adults working in nurseries and primary schools) and settings. Ongoing work to explore whether potential protective effects persist as community transmission patterns evolve, in particular in response to the emergence of new viral strains and the implementation of vaccination programmes, would also be beneficial.

## Conclusion

In a large occupational cohort, household exposure to young children was associated with a reduced risk of testing positive for SARS-CoV-2 and COVID-19 requiring hospitalisation (non-significant). Verification of this finding is needed in other populations and settings. Less equivocally, to date increased household exposure to young children has not been associated with an increased risk of COVID-19, even during periods where schools are open and there is active transmission of SARS-CoV2 in the community. These findings have potential for informing policy on nursery and school closure.

## Data Availability

Since our analysis involved data on unconsented participants, we are unable to share individual level data.

## Declaration of interests

AS is a co-owner of two nurseries which provide care to pre-school children.

## Acknowledgments

David McAllister is funded via a Wellcome Trust Intermediate Clinical Fellowship and Beit Fellowship (201492/Z/16/Z). Anoop Shah is funded via the British Heart Foundation through an Intermediate Clinical Research Fellowship (FS/19/17/34172).

## Data sharing statement

Data may be accessed via a secure platform following successful application to the Public Benefit and Privacy Panel via application to the electronic Data Research Information Services of Public Health Scotland.

## Approvals

This project was approved by the Public Benefit and Privacy Panel (2021-0013).

## Licence

The Corresponding Author has the right to grant on behalf of all authors and does grant on behalf of all authors, a worldwide licence (http://www.bmj.com/sites/default/files/BMJ%20Author%20Licence%20March%202013.doc) to the Publishers and its licensees in perpetuity, in all forms, formats and media (whether known now or created in the future), to i) publish, reproduce, distribute, display and store the Contribution, ii) translate the Contribution into other languages, create adaptations, reprints, include within collections and create summaries, extracts and/or, abstracts of the Contribution and convert or allow conversion into any format including without limitation audio, iii) create any other derivative work(s) based in whole or part on the on the Contribution, iv) to exploit all subsidiary rights to exploit all subsidiary rights that currently exist or as may exist in the future in the Contribution, v) the inclusion of electronic links from the Contribution to third party material where-ever it may be located; and, vi) licence any third party to do any or all of the above. All research articles will be made available on an open access basis (with authors being asked to pay an open access fee—see http://www.bmj.com/about-bmj/resources-authors/forms-policies-and-checklists/copyright-open-access-and-permission-reuse). The terms of such open access shall be governed by a Creative Commons licence—details as to which Creative Commons licence will apply to the research article are set out in our worldwide licence referred to above.

## Public and patient involvement

No patients were involved in setting the research question or the outcome measures, nor were they involved in developing plans for design of the study. No patients were asked to advise on interpretation or writing up of results

## What is already known on this topic

- Young children are less likely to develop COVID-19 and severe disease than adults, possibly in part due to cross-protective immunity to SARS-CoV-2.
- Whether contact with young children offers adults a degree of protection from COVID-19 or not is unknown.
- Two studies have suggested that contact with children may be inversely associated with COVID-19 risk (risk of infection, hospitalisation or death from COVID-19).

## What this study adds

- In our large cohort, adults with young children were at lower risk of testing positive for SARS-CoV-2 and possibly also of COVID-19 requiring hospitalisation.
- Adults living young children were not at increased risk of COVID-19, including during August to October 2020 when nurseries and schools were open for all children.
- Verification of this finding is warranted in other populations and settings.

## Notes

### Competing Interest Statement

AS owns two nursery schools.

### Funding Statement

David McAllister is funded via a Wellcome Trust Intermediate Clinical Fellowship and Beit Fellowship (201492/Z/16/Z) and Anoop Shah is funded via the British Heart Foundation through an Intermediate Clinical Research Fellowship (FS/19/17/34172). The funders had no role in the study design; in the collection, analysis, and interpretation of data; in the writing of the report; and in the decision to submit the article for publication

### Author Declarations

Scotland Public Benefit and Privacy Panel

### Summary of Updates

Added additional follow-up time (and hence events) including the period after schools re-opened.

