## Supplemental Tables for "Living with Children and Adults’ Risk of COVID-19: Observational Study"

### Table S1 Comorbidity among adults living in healthcare worker households by number of young children in household

|  | 0 children aged 0-11 | 1 child aged 0-11 | 2 children aged 0-11 | 3 or more children aged 0-11 |
| --- | --- | --- | --- | --- |
| Any comorbidity | 33470 (13.87) | 3883 (9.43) | 1859 (7.82) | 286 (7.43) |
| Ischaemic heart disease | 4067 (1.69) | 256 (0.62) | 74 (0.31) | 13 (0.34) |
| Other heart disease | 7298 (3.02) | 604 (1.47) | 250 (1.05) | 38 (0.99) |
| Other circulatory system diseases | 4386 (1.82) | 494 (1.20) | 237 (1.00) | 49 (1.27) |
| Advanced chronic kidney disease | 224 (0.09) | 30 (0.07) | 14 (0.06) | 0 |
| Asthma and chronic lower respiratory disease | 5311 (2.20) | 745 (1.81) | 394 (1.66) | 65 (1.69) |
| Neurological disorders | 1249 (0.52) | 161 (0.39) | 91 (0.38) | 11 (0.29) |
| Decompensated liver disease | 203 (0.08) | 13 (0.03) | 8 (0.03) | 0 |
| Any immunological condition | 217 (0.09) | 25 (0.06) | 13 (0.05) | 0 |
| Malignant Neoplasms | 7854 (3.26) | 819 (1.99) | 443 (1.86) | 52 (1.35) |
| Disorders of oesophagus, stomach and duodenum | 5707 (2.37) | 676 (1.64) | 325 (1.37) | 50 (1.30) |
| Diabetes, type 1 | 1617 (0.67) | 274 (0.67) | 160 (0.67) | 17 (0.44) |
| Diabetes, type 2 | 7660 (3.17) | 618 (1.50) | 205 (0.86) | 32 (0.83) |
| Diabetes, unknown type | 470 (0.19) | 57 (0.14) | 24 (0.10) | 6 (0.16) |

### Tables S2a to S2d Number of adults living in healthcare worker households according household composition

#### Table S2a Number of adults aged ≥18 by number of young children and children aged 12-17 in household

| **Number of children aged 12 to 17 in household** | **0 children aged 0-11** | **1 child aged 0-11** | **2 children aged 0-11** | **3 or more children aged 0-11** |
| --- | --- | --- | --- | --- |
| 0 aged 12-17 | 208714 (86.51) | 29855 (72.47) | 21470 (90.27) | 3559 (92.44) |
| 1 aged 12 to 17 | 24359 (10.10) | 9521 (23.11) | 2051 (8.62) | 215 (5.58) |
| 2 aged 12 to 17 | 7596 (3.15) | 1677 (4.07) | 244 (1.03) | 61 (1.58) |
| 3+ aged 12 to 17 | 597 (0.25) | 145 (0.35) | 18 (0.08) | 15 (0.39) |

Table S2b Number of adults aged ≥18 by number of young children and adults aged 18 or older in household

| **Number of adults aged 18 or older in household** | **0 children aged 0-11** | **1 child aged 0-11** | **2 children aged 0-11** | **3 or more children aged 0-11** |
| --- | --- | --- | --- | --- |
| 1 aged ≥18 | 39395 (16.33) | 5026 (12.20) | 2041 (8.58) | 365 (9.48) |
| 2 aged ≥18 | 78980 (32.74) | 23042 (55.93) | 17499 (73.58) | 3273 (85.01) |
| 3+ aged ≥18 | 122891 (50.94) | 13130 (31.87) | 4243 (17.84) | 212 (5.51) |

#### Table S2c Number of adults aged ≥18 by number of young children and adults aged 65 to 74 in household

| **Number of adults aged 65 to 74 in household** | **0 children aged 0-11** | **1 child aged 0-11** | **2 children aged 0-11** | **3 or more children aged 0-11** |
| --- | --- | --- | --- | --- |
| 0 aged 65-74 | 219469 (90.97) | 39659 (96.26) | 23208 (97.58) | 3797 (98.62) |
| 1 aged 65-74 | 17701 (7.34) | 1310 (3.18) | 509 (2.14) | 43 (1.12) |
| 2 aged 65-74 | 3925 (1.63) | 229 (0.56) | 66 (0.28) | 10 (0.26) |
| 3+ aged 65-74 | 171 (0.07) | 0 | 0 | 0 |

#### Table S2d Number of adults aged ≥18 by number of young children and adults aged 75 or older in household

| **Number of adults aged 75 or older in household** | **0 children aged 0-11** | **1 child aged 0-11** | **2 children aged 0-11** | **3 or more children aged 0-11** |
| --- | --- | --- | --- | --- |
| 0 aged ≥75 | 231223 (95.84) | 40226 (97.64) | 23387 (98.33) | 3817 (99.14) |
| 1 aged ≥75 | 8290 (3.44) | 827 (2.01) | 361 (1.52) | 17 (0.44) |
| 2 aged ≥75 | 1674 (0.69) | 145 (0.35) | 35 (0.15) | 16 (0.42) |
| 3+ aged ≥75 | 79 (0.03) | 0 | 0 | 0 |

### Table S3 Risk and hazard ratios for COVID-19 requiring hospitalisation for adults living in patient-facing healthcare worker households* by number of young children in household.

|  | **0 children aged 0-11** | **1 child aged 0-11** | **2 children aged 0-11** | **3 or more children aged 0-11** | **Per child** |
| --- | --- | --- | --- | --- | --- |
| Events | 272 | 41 | 17 | 1 | - |
| N | 137697 | 25461 | 15485 | 2627 | - |
| Risk per 10,000 | 19.8 | 16.1 | 11.0 | 3.8 | - |
| Unadjusted | 1 | 0.82 (0.58-1.14) | 0.57 (0.35-0.93) | 0.20 (0.03-1.41) | 0.75 (0.62-0.90) |
| Model 1 | 1 | 0.94 (0.67-1.32) | 0.67 (0.41-1.12) | 0.24 (0.03-1.69) | 0.82 (0.68-0.99) |
| Model 2 | 1 | 0.97 (0.69-1.37) | 0.74 (0.44-1.24) | 0.26 (0.04-1.84) | 0.85 (0.71-1.03) |
| Model 3 | 1 | 0.99 (0.70-1.39) | 0.75 (0.45-1.25) | 0.26 (0.04-1.87) | 0.86 (0.71-1.03) |
| Model 4 | 1 | 1.03 (0.73-1.45) | 0.82 (0.49-1.37) | 0.29 (0.04-2.07) | 0.89 (0.74-1.08) |

Model fitting and covariates as per footnote of Table 2 in the main manuscript. * Households where at least one healthcare worker occupies a patient facing role.

Figure S1 – Causal diagram showing assumed role of covariates included in model 4 (Tables 2-4 of main manuscript)

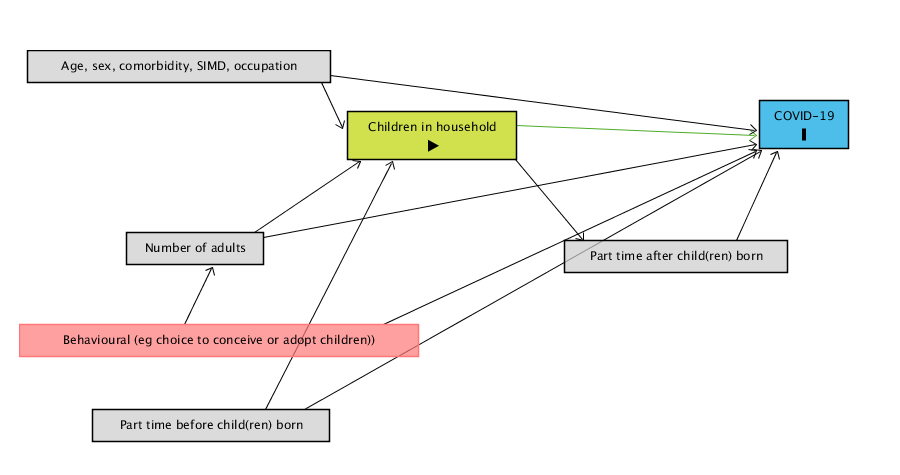

Green box indicates exposure, blue box outcome, grey boxes indicate variable/groups of variables conditioned on.

### Table S4 Hazard ratios for COVID-19 requiring hospitalisation for adults living in healthcare worker households by number of persons in household of different ages

|  | **Per child  aged 0 to 4** | **Per child  aged 5 to 11** | **Per child  aged 12 to 17** | **Per adult  aged 18 or above** |
| --- | --- | --- | --- | --- |
| Unadjusted | 0.63 (0.46-0.88) | 1.03 (0.84-1.26) | 0.91 (0.84-0.99) | 0.84 (0.70-1.02) |
| Model 1 | 0.84 (0.61-1.15) | 1.02 (0.84-1.25) | 0.93 (0.85-1.01) | 0.91 (0.74-1.11) |
| Model 2 | 0.84 (0.61-1.16) | 1.04 (0.85-1.28) | 1.00 (0.91-1.09) | 0.92 (0.75-1.13) |
| Model 3 | 0.85 (0.62-1.17) | 1.05 (0.86-1.29) | 1.00 (0.92-1.09) | 0.93 (0.76-1.14) |
| Model 4 | 0.88 (0.64-1.22) | 1.07 (0.87-1.30) | 1.00 (0.91-1.09) | 0.96 (0.78-1.18) |

Model specification and covariates as per footnote of Table 4.

### Table S5 Risks and hazard ratios for severe COVID-19 for adults living in healthcare worker households by number of young children in household

|  | **0 children aged 0-11** | **1 child aged 0-11** | **2 children aged 0-11** | **3 or more children aged 0-11** | **Per child** |
| --- | --- | --- | --- | --- | --- |
| N adults with severe COVID-19 |  |  |  |  | - |
| Total N adults | 241266 | 41198 | 23783 | 3850 | - |
| Risk per 10,000 | 3.6 | 2.7 | 1.7 | 0.0 | - |
| Unadjusted | 1 | 0.75 (0.40-1.40) | 0.48 (0.18-1.30) | - | 0.68 (0.47-0.98) |
| Model 1 | 1 | 1.17 (0.62-2.23) | 0.88 (0.32-2.37) | - | 0.94 (0.66-1.33) |
| Model 2 | 1 | 1.25 (0.67-2.34) | 0.87 (0.31-2.41) | - | 0.95 (0.68-1.33) |
| Model 3 | 1 | 1.29 (0.69-2.42) | 0.86 (0.31-2.41) | - | 0.93 (0.67-1.30) |
| Model 4 | 1 | 1.36 (0.73-2.56) | 0.97 (0.34-2.74) | - | 1.01 (0.72-1.42) |

Model fitting and covariates as per footnote of Table 2 in the main manuscript.

### Table S6 Hazard ratios for severe COVID-19 for adults living in healthcare worker households by number of persons in household of different ages

|  | Per child  aged 0 to 5 | Per child  aged 6 to 11 | Per child  aged 12 to 17 | Per adult  aged 18 or above |
| --- | --- | --- | --- | --- |
| Unadjusted | 0.57 (0.23-1.39) | 0.54 (0.32-0.93) | 1.22 (1.04-1.43) | 0.74 (0.48-1.13) |
| Model 1 | 0.96 (0.41-2.22) | 0.59 (0.35-1.00) | 1.21 (1.02-1.44) | 0.93 (0.61-1.43) |
| Model 2 | 0.90 (0.38-2.15) | 0.59 (0.35-1.02) | 1.20 (0.99-1.45) | 0.97 (0.64-1.46) |
| Model 3 | 0.87 (0.36-2.08) | 0.60 (0.35-1.02) | 1.21 (1.01-1.45) | 0.96 (0.64-1.45) |
| Model 4 | 0.95 (0.41-2.22) | 0.60 (0.35-1.04) | 1.21 (1.01-1.45) | 1.04 (0.69-1.57) |

Model specification and covariates as per footnote of Table 4.

### Scottish household survey

As, we were unable to find any published studies providing empirical evidence to support or refute the claim that adults who shared a household with young children have restricted activity outside the home, we downloaded and analysed the 2018 Scottish Household Survey (which is available via the UK data Service at <https://beta.ukdataservice.ac.uk/datacatalogue/series/series?id=2000048>).

The 2018 Scottish Household Survey includes a travel diary where a “random adult” respondent in each household is asked to record all the activities they travelled to/from within the previous 24 hours. These activities are then assigned to standard categories. The Scottish Household Survey also records the number of children aged 0-11 and the number of children aged 0-4 in each household. This therefore provides a high-quality contemporaneous dataset for comparing the activities of households containing different numbers of children.

We restricted the data to households where the “random adult” was aged 25 to 44. We did so to avoid confounding without having to resort to regression modelling. Because we were interested solely in relative differences, we did not use the survey weighting variables which account for oversampling of different geographic regions. We compared the activities using both the original categorisations from the Scottish Household Survey and after collapsing these into broader categories relevant to SARS-CoV-2 exposure. In the supplementary appendix we provide both tables in full; below for convenience we have reproduced the collapsed-categories as well as selected rows showing the original categorisations.

### Table S7 Collapsed categories of activities. Number (%) of households where the “random adult” travelled to/from activities according to the number of children aged 0 to 11.

|  | 0 | 1 | 2 | 3 or 4 |
| --- | --- | --- | --- | --- |
| Number of households | 1085 | 458 | 421 | 80 |
| *Age random adult in household - mean (standard deviation)* | 33.8 (5.9) | 35.7 (5.2) | 35.8 (5) | 35.05 (4.8%) |
| *Community activities, indoor* | 362 (33.4%) | 156 (34.1%) | 147 (34.9%) | 30 (37.5%) |
| *Community activities, outdoor* | 5 (0.5%) | 7 (1.5%) | 6 (1.4%) | 2 (2.5%) |
| *Community activities, indoor or outdoor* | 41 (3.8%) | 18 (3.9%) | 18 (4.3%) | 5 (6.3%) |
| *Educational* | 33 (3%) | 60 (13.1%) | 79 (18.8%) | 23 (28.8%) |
| *Seeing friends* | 89 (8.2%) | 21 (4.6%) | 24 (5.7%) | 1 (1.3%) |
| *Home (journeying to or from)* | 210 (19.4%) | 92 (20.1%) | 83 (19.7%) | 24 (30%) |
| *Hospital* | 11 (1%) | 6 (1.3%) | 5 (1.2%) | 1 (1.3%) |
| *Other* | 141 (13%) | 77 (16.8%) | 69 (16.4%) | 14 (17.5%) |
| *Private* | 72 (6.6%) | 19 (4.1%) | 21 (5%) | 8 (10%) |
| *Relatives* | 106 (9.8%) | 38 (8.3%) | 30 (7.1%) | 7 (8.8%) |
| *Work* | 516 (47.6%) | 192 (41.9%) | 171 (40.6%) | 28 (35%) |

### Table S8 Collapsed categories of activities. Number (%) of households where the “random adult” travelled to/from activities according to the number of children aged 0 to 4

|  | 0 | 1 | 2 or 3 |
| --- | --- | --- | --- |
| *Number of households* | 1493 | 432 | 119 |
| *Age random adult in household - mean (standard deviation)* | 34.9 (5.8) | 34.3 (4.9) | 33.8 (4.9) |
| *Community activities, indoor* | 504 (33.8%) | 147 (34%) | 44 (37%) |
| *Community activities, outdoor* | 9 (0.6%) | 6 (1.4%) | 5 (4.2%) |
| *Community activities, indoor or outdoor* | 60 (4%) | 19 (4.4%) | 3 (2.5%) |
| *Educational* | 102 (6.8%) | 64 (14.8%) | 29 (24.4%) |
| *Seeing friends* | 107 (7.2%) | 22 (5.1%) | 6 (5%) |
| *Home (journeying to or from)* | 291 (19.5%) | 88 (20.4%) | 30 (25.2%) |
| *Hospital* | 15 (1%) | 6 (1.4%) | 2 (1.7%) |
| *Other* | 208 (13.9%) | 81 (18.8%) | 12 (10.1%) |
| *Private* | 94 (6.3%) | 15 (3.5%) | 11 (9.2%) |
| *Relatives* | 135 (9%) | 36 (8.3%) | 10 (8.4%) |
| *Work* | 694 (46.5%) | 170 (39.4%) | 43 (36.1%) |

### Table S9 Number (%) of households where the “random adult” travelled to/from activities according to the number of children aged 0 to 11

| **Children aged 0 to 11** | **0** | **1** | **2** | **3or4** |
| --- | --- | --- | --- | --- |
| **Number of households** | 1085 | 458 | 421 | 80 |
| **Age random adult in HH - mean (sd)** | 33.8 (5.9) | 35.7 (5.2) | 35.8 (5) | 35.05 (4.8) |
| **Other medical related eg. dentist, physio** | 5 (0.5%) | 7 (1.5%) | 1 (0.2%) | 1 (1.3%) |
| **Place of worship eg. church, mosque** | 5 (0.5%) | 8 (1.7%) | 5 (1.2%) | 1 (1.3%) |
| **The bank** | 2 (0.2%) | 1 (0.2%) | 1 (0.2%) | 0 (0%) |
| **The cinema** | 8 (0.7%) | 5 (1.1%) | 2 (0.5%) | 1 (1.3%) |
| **The Council offices** | 2 (0.2%) | 1 (0.2%) | 0 (0%) | 0 (0%) |
| **The Doctors** | 6 (0.6%) | 6 (1.3%) | 5 (1.2%) | 2 (2.5%) |
| **The gym** | 20 (1.8%) | 12 (2.6%) | 12 (2.9%) | 3 (3.8%) |
| **The pub** | 22 (2%) | 3 (0.7%) | 0 (0%) | 2 (2.5%) |
| **The restaurant** | 31 (2.9%) | 13 (2.8%) | 9 (2.1%) | 1 (1.3%) |
| **The shops** | 287 (26.5%) | 119 (26%) | 130 (30.9%) | 24 (30%) |
| **The park** | 5 (0.5%) | 7 (1.5%) | 6 (1.4%) | 2 (2.5%) |
| **Another entertainment/public activity** | 15 (1.4%) | 10 (2.2%) | 6 (1.4%) | 2 (2.5%) |
| **Participating in sport/exercise** | 26 (2.4%) | 9 (2%) | 12 (2.9%) | 3 (3.8%) |
| **College** | 0 (0%) | 4 (0.9%) | 1 (0.2%) | 0 (0%) |
| **School** | 0 (0%) | 1 (0.2%) | 1 (0.2%) | 1 (1.3%) |
| **School/college/university** | 24 (2.2%) | 5 (1.1%) | 2 (0.5%) | 0 (0%) |
| **School/nursery** | 8 (0.7%) | 51 (11.1%) | 77 (18.3%) | 22 (27.5%) |
| **University** | 1 (0.1%) | 1 (0.2%) | 0 (0%) | 0 (0%) |
| **Your friends** | 89 (8.2%) | 21 (4.6%) | 24 (5.7%) | 1 (1.3%) |
| **Home** | 210 (19.4%) | 92 (20.1%) | 83 (19.7%) | 24 (30%) |
| **The Hospital** | 11 (1%) | 6 (1.3%) | 5 (1.2%) | 1 (1.3%) |
| **Another personal appointment** | 43 (4%) | 21 (4.6%) | 18 (4.3%) | 4 (5%) |
| **Escort - other** | 12 (1.1%) | 22 (4.8%) | 23 (5.5%) | 4 (5%) |
| **On holiday** | 2 (0.2%) | 0 (0%) | 0 (0%) | 0 (0%) |
| **Other** | 24 (2.2%) | 6 (1.3%) | 7 (1.7%) | 3 (3.8%) |
| **Volunteering/caring** | 8 (0.7%) | 4 (0.9%) | 2 (0.5%) | 2 (2.5%) |
| **Your day trip** | 13 (1.2%) | 7 (1.5%) | 16 (3.8%) | 1 (1.3%) |
| **Your meeting** | 46 (4.2%) | 20 (4.4%) | 13 (3.1%) | 2 (2.5%) |
| **Take the dog out** | 42 (3.9%) | 5 (1.1%) | 10 (2.4%) | 4 (5%) |
| **Your cycle** | 2 (0.2%) | 2 (0.4%) | 0 (0%) | 0 (0%) |
| **Your drive** | 3 (0.3%) | 0 (0%) | 1 (0.2%) | 0 (0%) |
| **Your run** | 5 (0.5%) | 3 (0.7%) | 5 (1.2%) | 1 (1.3%) |
| **Your walk** | 21 (1.9%) | 10 (2.2%) | 6 (1.4%) | 3 (3.8%) |
| **Your relatives** | 106 (9.8%) | 38 (8.3%) | 30 (7.1%) | 7 (8.8%) |
| **Work** | 516 (47.6%) | 192 (41.9%) | 171 (40.6%) | 28 (35%) |

### Table S10 Number (%) of households where the “random adult” travelled to/from activities according to the number of children aged 0 to 4

| **Children aged 0 to 4** | **0** | **1** | **2or3** |
| --- | --- | --- | --- |
| **Number of households** | 1493 | 432 | 119 |
| **Age random adult in HH - mean (sd)** | 34.9 (5.8) | 34.3 (4.9) | 33.8 (4.9%) |
| **Other medical related eg. dentist, physio** | 9 (0.6%) | 4 (0.9%) | 1 (0.8%) |
| **Place of worship eg. church, mosque** | 10 (0.7%) | 9 (2.1%) | 0 (0%) |
| **The bank** | 4 (0.3%) | 0 (0%) | 0 (0%) |
| **The cinema** | 14 (0.9%) | 2 (0.5%) | 0 (0%) |
| **The Council offices** | 3 (0.2%) | 0 (0%) | 0 (0%) |
| **The Doctors** | 14 (0.9%) | 5 (1.2%) | 0 (0%) |
| **The gym** | 32 (2.1%) | 11 (2.5%) | 4 (3.4%) |
| **The pub** | 24 (1.6%) | 1 (0.2%) | 2 (1.7%) |
| **The restaurant** | 40 (2.7%) | 10 (2.3%) | 4 (3.4%) |
| **The shops** | 398 (26.7%) | 124 (28.7%) | 38 (31.9%) |
| **The park** | 9 (0.6%) | 6 (1.4%) | 5 (4.2%) |
| **Another entertainment/public activity** | 21 (1.4%) | 10 (2.3%) | 2 (1.7%) |
| **Participating in sport/exercise** | 39 (2.6%) | 10 (2.3%) | 1 (0.8%) |
| **College** | 3 (0.2%) | 2 (0.5%) | 0 (0%) |
| **School** | 2 (0.1%) | 1 (0.2%) | 0 (0%) |
| **School/college/university** | 28 (1.9%) | 2 (0.5%) | 1 (0.8%) |
| **School/nursery** | 69 (4.6%) | 60 (13.9%) | 29 (24.4%) |
| **University** | 2 (0.1%) | 0 (0%) | 0 (0%) |
| **Your friends** | 107 (7.2%) | 22 (5.1%) | 6 (5%) |
| **Home** | 291 (19.5%) | 88 (20.4%) | 30 (25.2%) |
| **The Hospital** | 15 (1%) | 6 (1.4%) | 2 (1.7%) |
| **Another personal appointment** | 62 (4.2%) | 23 (5.3%) | 1 (0.8%) |
| **Escort - other** | 36 (2.4%) | 22 (5.1%) | 3 (2.5%) |
| **On holiday** | 2 (0.1%) | 0 (0%) | 0 (0%) |
| **Other** | 28 (1.9%) | 10 (2.3%) | 2 (1.7%) |
| **Volunteering/caring** | 11 (0.7%) | 2 (0.5%) | 3 (2.5%) |
| **Your day trip** | 24 (1.6%) | 12 (2.8%) | 1 (0.8%) |
| **Your meeting** | 58 (3.9%) | 20 (4.6%) | 3 (2.5%) |
| **Take the dog out** | 54 (3.6%) | 3 (0.7%) | 4 (3.4%) |
| **Your cycle** | 4 (0.3%) | 0 (0%) | 0 (0%) |
| **Your drive** | 3 (0.2%) | 0 (0%) | 1 (0.8%) |
| **Your run** | 11 (0.7%) | 3 (0.7%) | 0 (0%) |
| **Your walk** | 25 (1.7%) | 9 (2.1%) | 6 (5%) |
| **Your relatives** | 135 (9%) | 36 (8.3%) | 10 (8.4%) |
| **Work** | 694 (46.5%) | 170 (39.4%) | 43 (36.1%) |
